# Emergence of macrolide-resistant *Bordetella pertussis* in France, 2024: out of China

**DOI:** 10.1101/2025.03.15.25324024

**Authors:** Valérie Bouchez, Noémie Lefrancq, Julie Toubiana, Carla Rodrigues, Sylvain Brisse

## Abstract

**Objectives:** Pertussis is a highly contagious, potentially fatal vaccine-preventable respiratory disease, primarily caused by *Bordetella pertussis* (*Bp*). Macrolides constitute the first-line treatment for pertussis, reducing bacterial carriage and transmission. Despite consistent surveillance, only one resistant isolate had ever been reported in France before 2024 (in 2011). Here, we report 14 macrolide-resistant *Bp* (MRBP) cases, collected in France between February and November 2024, during the largest whooping cough outbreak of the last 20 years. We aimed to investigate whether these MRBP arose from macrolide-susceptible *Bp* (MSBP) in France or were instead imported.

**Methods:** Illumina sequencing was performed for all French isolates from 2024 and in addition, long-read sequencing was performed for the MRBP isolates. We compared the 14 MRBP genomic sequences with 1,571 macrolide-susceptible cultivated *B. pertussis* isolates collected in France (1993-2024, including 331 from 2024), the MRBP isolate from 2011, and 824 *Bp* collected in China (2018-2024), including 596 MRBP (75.2%).

**Results:** Phylogenetic analysis based on whole-genome single nucleotide polymorphisms revealed that the French MRBP belong to three separate branches nested within the diversity of MRBP isolates from China, suggesting three independent introductions into France. Two of these branches comprised more than one isolate, detected across several French administrative regions, indicating forward transmission and spatial dissemination. The MRBP isolates from France and China belonged to a single clade of the *ptxP*3 lineage corresponding to the previous genotype denomination MT28.

**Conclusion:** The rise of MRBP in France is driven by importation followed by local dissemination. This exceptional emergence is concerning, given the high expected fitness of PRN-negative *ptxP3* MRBP isolates in acellular vaccination countries. Besides vaccination, effective control of MRBP will require enhanced surveillance, strict adherence to transmission control guidelines, and prudent use of macrolides to avoid selective pressure favouring MRBP.

## Introduction

Pertussis is a highly contagious vaccine-preventable respiratory disease, primarily caused by *Bordetella pertussis* (*Bp*), that can be lethal in young infants. In 2024, an intense resurgence of pertussis occurred in Europe and elsewhere (1), after circulating at very low levels since 2020. In France, an increase in the incidence of pertussis was reported (1,2).

Macrolides constitute the first-line treatment for pertussis, reducing bacterial carriage and transmission. Of major public health concern, macrolide-resistant *Bp* (MRBP) isolates have been reported in China since 2011, now reaching > 90% in some settings (4–7) . Elsewhere, only sporadic MRBP cases have been reported up to now, including one case in France in 2011 (8). In China, MRBP isolates belong to two main phylogenetic clades called *ptxP1* and *ptxP3,* which are marked by a mutation in the promoter of the pertussis toxin (*ptx*P) gene cluster (4,6). Whereas *ptxP1* MRBP isolates emerged around 2010 in China, *ptxP3* MRBP emerged around 2016 and became predominant in some areas, such as Shanghai (6,9,10). This shift in *ptxP* genotype prevalence that was observed since 2019 in China might be associated with changes in pertussis vaccines in the country, which occurred approximately since 1995 from whole cell vaccines (wP) to acellular vaccines (aP) (11). *ptxP3* isolates, particularly those that have lost the expression of the vaccine antigen pertactin (PRN), were previously inferred to have a much higher fitness, and thus to spread more rapidly, in aP-using countries (12).

Here, we report 14 MRBP isolates from France collected between February and November 2024. Through genome-wide phylogenetic comparisons, we investigated whether the MRBPs arose from macrolide-susceptible *Bp* (MSBP) in France or were imported from abroad. We discuss possible reasons for their successful dissemination in France in light of their genomic features.

## Methods

*Bp* isolates were collected through the Reseau Microbiologique de la Coqueluche (REMICOQ) network, which comprises laboratories from the Reseau National de la Coqueluche (RENACOQ) paediatric hospital-based surveillance network and additional collaborative hospital and outpatient laboratories (1). Culture was attempted at the French National Reference Center for Whooping Cough (NRC). Microbiological identification and characterisation were performed as previously described (1,13). Antibiotic susceptibility testing was performed by disk diffusion for ampicillin, cephalexin, streptomycin, trimethoprim-sulfamethoxazole, erythromycin, azithromycin, clarithromycin and spiramycin as previously described (1) and macrolide resistance was confirmed by E-test. In cases where macrolide resistance was confirmed, an extended panel of antibiotics was tested by disk diffusion, including piperacillin-tazobactam, ceftazidime, ceftriaxone, imipenem, doxycycline, amikacin, tobramycin, and tetracycline. We sequenced all *Bp* isolates from France in 2024 (see Supplementary appendix) and compared the genomic sequences of the 14 MRBP isolates with (i) 1,572 *Bp* collected in France (1993-2024, including the MRBP isolate from 2011 and 331 MSBP isolates sequenced in 2024), and (ii) 824 *Bp* collected in China in two large studies with publicly available genomic data (2018-2024; NCBI BioProjects PRJNA908268 and PRJNA1071282), including 596 MRBP (75.2%) (4,6). Phylogenetic analysis was performed using whole-genome single nucleotide polymorphisms (SNP) as previously described (12). Briefly, the SNP-based alignment was used to reconstruct the phylogenetic relationships of the isolates, using IQ-tree v2.3.6 and a GTR+F substitution model. The figures were generated using ggtree v3.2. Additionally, Nanopore sequencing was performed on the 14 French MRBP to define the number of mutated copies of the *23S rRNA* gene (see Supplementary appendix).

Finally, in response to the emergence of MRBP in France, we adapted a qPCR assay (14) to detect MRBP directly from respiratory samples (see Supplementary Appendix, protocol posted on protocols.io (14)). Using this method, we screened the 534 pertussis-toxin positive samples (as defined by a positive qPCR for pertussis toxin gene *ptxA*) collected in 2024.

## Results

Out of 345 *Bp* isolates collected across France in 2024, 14 MRBP were identified. These were collected from 4 infants under 12 months of age, 5 children aged 1 to 6 years, and 5 adults ranging from 18 to 79 years old. Of the 9 documented cases, 2 required hospitalisation, including one newborn admitted to intensive care. The 14 isolates produced pertussis toxin (PT), filamentous haemagglutinin (FHA) and the fimbriae of serotype FIM2. Whereas one isolate was PRN-positive (see branch I3, Figure 1B), the 13 others were PRN-negative (branches I1 and I2). This contrasted with the macrolide-susceptible *Bp* (MSBP) isolates collected over the study period, which all produced PRN except one. The 14 MRBP isolates carried the A2047G mutation in the 3 copies of the *23S rRNA* gene, consistent with their antimicrobial resistance phenotype. Based on antigen genes (15), the 14 MRBP isolates belonged to the same genotype, Bp-agST4, and were characterised by a *ptxP3* promoter. Differently, among MSBP isolates from 2024, 18 were characterised by a *ptxP1* promoter and 11 or these displayed a new allele for *fim3* (*fim3-26,* characterised by a synonymous mutation C87T, also observed in *fim3*-4, and an additional non-synonymous mutation leading to amino-acid substitution V165D).

**Figure 1.**
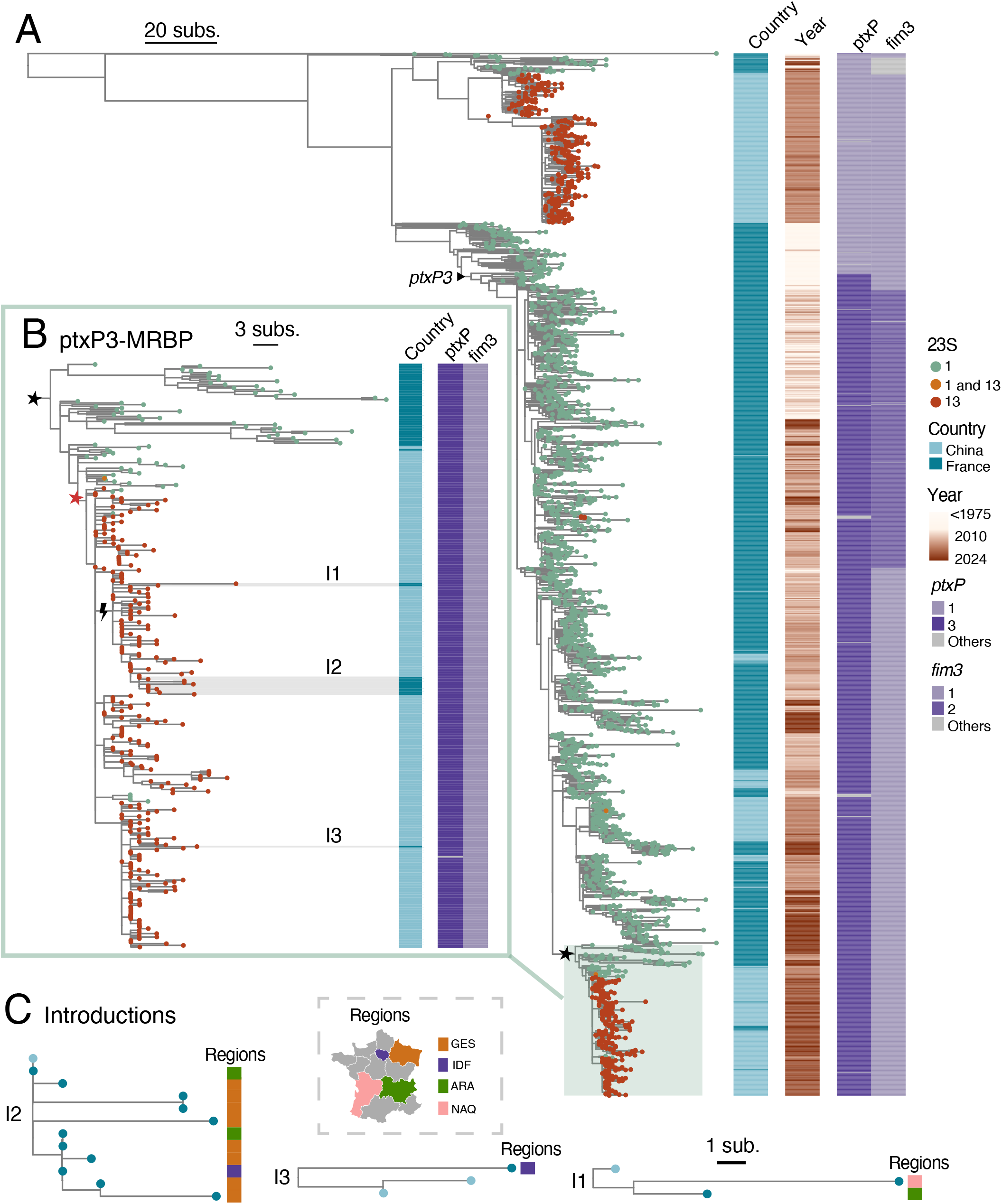
Phylogenetic diversity and geographic dissemination of macrolide-resistant Bordetella pertussis, France 2024. The tree was constructed based on whole genome SNPs using method described in Lefrancq *et al*. **Panel A** presents the tree obtained from the whole dataset: tree leaves are red for MRBP, (allele 13, *i.e.*, 23S rRNA A2047G mutation) or green for MSBP (allele 1); 2 isolates with a single mutated copy of the *23S rRNA* gene are colored in orange. Landmark mutation *ptxP3* is indicated at corresponding node with a black triangle. The columns indicate the country of origin (France: dark blue; China: light blue); years of collection (from 1975 to 2024), *ptxP* and *fim3* alleles (light purple for allele 1, darker purple for alleles *fim3*-2 and *ptxP3*, light grey for other or undefined alleles). The scales indicate the number of SNPs per genome. **Panel B** focuses on the emerging *ptxP3*-MRBP clade (349 isolates). Evolved SNPs shared by this clade at the black star location are detailed in the Supplementary Appendix, Table S2; the 23S rRNA mutation is indicated by a red star; a pertactin-inactivating mutation by a lightning symbol. The pertactin-negative subclade was introduced twice into France. The three introductions of MRBP into France are labelled as I1, I2 and I3 and highlighted by light grey horizontal bars (see Table S3 for details). **Panel C** presents the detailed trees of the three introductions of MRBP into France, and their geographic origins, as indicated in the map colored according to French administrative regions: orange for ‘Grand Est’ (GES), purple for ‘Ile de France’ (IDF), green for ‘Auvergne Rhône Alpes’ (ARA) and salmon for ‘Nouvelle Aquitaine’ (NAQ). Circles at branch tips are colored according to country (light blue: China; darker blue: France). The tree can be interactively visualized in Microreact (**https://microreact.org/project/5mvNLVqAoPoT2V6y2kUPFw-bp-cnr-china-all).**

Phylogenetic analysis based on whole-genome single nucleotide polymorphisms (Figure 1) revealed that the 14 MRBP belong, together with isolates from China, to a cluster of 349 MRBP isolates (Figure 1B; the *ptxP3*-MRBP clade). However, they fall into three separate branches nested within the diversity of isolates from China, indicating three independent introductions of MRBP into France. Two of these branches comprised more than one isolate (branches I2 and I1, with 11 and 2 isolates, respectively; table S2), detected across several French administrative regions, suggesting forward transmission and spatial dissemination (Figure 1C).

Eight mutations were found in isolates of the *ptxP3*-MRBP clade and may have occurred in a common ancestor preceding this lineage’s expansion (see black star in Figure 1; Table 1). The first corresponds to the A2047G mutation within the *23S rRNA* gene. Two others are observed within the *prn* gene (C531T, leading to allele 150) and within a putative toxin gene (T198M in gene BP1251). Two other non-synonymous SNPs are located in genes involved in regulatory functions: V67L in BP0983 coding for a transcriptional regulator controlling *mexAB-oprM* operon (16) and A165V in BP1924 coding for a TetR/AcrR transcriptional regulator. A further non-synonymous mutation (leading to L213P) is located in gene BP2754 coding for a penicillin-binding protein. The two remaining mutations were localised in hypothetical or miscellaneous proteins.

**Table 1.** Specific non-synonymous mutations observed in the *ptxP3*-MRBP clade.

| SNP position | Gene-name | Gene-id | Gene-category | SNP | Localization | Product | Nb isolates |
| --- | --- | --- | --- | --- | --- | --- | --- |
| 490040 | BP_RS02415 | BP0478 | Miscellaneous | G81A | cytoplasmic | HAD family hydrolase | 346 |
| 1021456 | BP_RS04885 | BP0983 | Regulation | V67L | cytoplasmic | LysR family transcriptional regulator | 349 |
| 1317235 | BP_RS06250 | BP1251 | Virulence | T198M | unknown | Putative toxin | 349 |
| 2026485 | BP_RS09590 | BP1924 | Regulation | A165V | cytoplasmic | TetR.AcrR transcriptional regulator | 349 |
| 2641711 | BP_RS12490 | BP2494 | Hypothetical | D165N | cytoplasmic | VMA domain-containing protein | 349 |
| 2928270 | pbpC | BP2754 | Cell surface | L213P | cytoplasmic | Penicillin-binding protein 1C | 349 |
| 1098621 | prn | BP1054 | Virulence | C531T | membrane | autotransporter | 264 |
| 2153195 | 23S-rRNA | Bpr02/Bpr05/Bpr08 | Ribosomal | A2047G | ribosomal | ribosomal RNA | 267 |

SNP, single nucleotide polymorphism

Regarding the screening of respiratory samples using the in-house qPCR protocol (17), out of 248 eligible samples based on *ptxA* qPCR Ct value, 3 (1.2%) were classified as MRBP. Culture attempts for these 3 samples were unsuccessful. Overall, combining isolates and respiratory samples, MRBP accounted for 2.8% (17/593) of the French 2024 tests.

## Discussion

After four years (2020-2023) with low-level circulation of *Bp*, a large epidemic of whooping cough occurred in 2024 in France and other countries (1). Whereas a resurgence was expected, the emergence of MRBP was not. Only one MRBP isolate had been previously reported in France, in 2011 (18). Differently, in China, the first MRBP isolates were reported in 2011 in Shandong province, and increased in frequency in subsequent years, reaching more than 90% in some areas such as Shanghai (3,6,9,10).

Whereas the first MRBP reported from China were characterised by a *ptxP1* and *fhaB3* genotype, those identified since 2016 are mostly of the *ptxP3* and *fhaB1* genotype (4,5,7,10), and before 2024 the expansion of this particularly fast-emerging MRBP genotype was documented exclusively in China. In France, most circulating MSBP isolates since the introduction of vaccination are *ptxP3* and *fhaB1* (1,12,13,19). The only MRBP isolate from France collected before 2024 (in 2011) was of genotype *ptxP21* and *fhaB1*. The emergence of *ptxP3* and *fhaB1* MRBP isolates in France in 2024 thus raised the question of their origin: have they been introduced in the country from China or elsewhere, or are they due to a local evolution from MSBP isolates in France?

While our data suggest three direct introductions from China to France, cryptic transmission through intermediary unsampled countries cannot be excluded. More globally representative sampling and genomic analyses are needed to shed light on *Bp* global spread. Understanding the impact of global mobility versus local transmission on the emergence of MRBP is crucial for defining effective strategies to control macrolide-resistant pertussis. Although they emerged 15 years ago in China, MRBP are only emerging in France in 2024, despite disease activity in 2010-2019 (1). As MRBP initially emerged in China within the *ptxP1* branch, we propose that this surprising 15-year delay might be due to the lower fitness of *ptxP1* isolates in acellular vaccination countries, compared to *ptxP3* isolates. The *ptxP3*-MRBP clade emerged in China only around 2016 and became predominant after 2020 (4,6). Besides, the loss of PRN expression provides a further fitness gain, particularly in the *fim3*-2 subbranch of *ptxP3* (12). The dissemination in France of the *ptxP*3, *fim3*-2, PRN-negative MRBP genotype is consistent with its high predicted fitness. Such MRBP may therefore quickly emerge in other countries too. A first PRN-negative *ptxP3*-MRBP was also reported in Finland in 2024 (20). The use of macrolides, and how it differs between countries, might be a factor favouring the transmission of this 23S rRNA A2047G-carrying genotype, relative to MSBP. Whether some of the 7 other mutations shared by these emerging MRBP isolates also contribute to an increased fitness remains to be defined.

The rise of MRBP in France in 2024 is exceptional and concerning. Effective control of macrolide resistance will require novel diagnostics, enhanced surveillance and strict adherence to transmission control guidelines. Macrolides are the drug of choice for treating other respiratory pathogens including *Mycoplasma pneumoniae,* and macrolide resistance is also reported for this species (21). To slow the rise of MRBP, prudent use of macrolides to treat pertussis and other infections or to limit their spread should be recommended to avoid selective pressure favouring resistant isolates. Besides, defining effective alternative treatments, particularly for newborns, is essential for the clinical management of future macrolide-resistant cases.

## Data Availability

Raw FASTQ data from French isolates are available in the European Nucleotide Archive (https://www.ebi.ac.uk/ena; Projects: PRJEB21744 and PRJEB42353). The qPCR protocol for detection of macrolide-resistant Bordetella pertudssis from clinical samples was posted on protocols.io at https://dx.doi.org/1017504/protocols.io.kqdg3q4y1v25/v1

## Acknowledgements

We thank the Institut Pasteur’s Mutualized Platform for Microbiology (P2M) for genomic sequencing. We acknowledge support to the French whooping cough surveillance from colleagues of the REMICOQ network and Public Health France (Santé Publique France). Nathalie Armatys, Annie Landier and Julien Cordani are acknowledged for conducting the microbiological screening and characterization. Nora Zidane performed the ONT sequencing of MRBP strains.

## Author contributions

CR and SB supervised the collection of isolates and data, with input from JT. VB and NL performed the genomic analyses, with input from CR and SB. VB, CR and SB wrote the manuscript, which was approved by all authors.

## Transparency declaration

The authors declare that they have no conflicts of interest.

