## Supplementary Appendix for "Emergence of macrolide-resistant *Bordetella pertussis* in France, 2024: out of China"

**Table of Contents:**

Supplementary Methods	2

Supplementary Tables	5

Supplementary References	7

**Supplementary Methods**

Vaccine antigen production

Vaccine antigen production was assessed using western-blot for pertactin (PRN), pertussis toxin (PT) and filamentous haemagglutinin (FHA) and using agglutination for FIM2 and FIM3 serotyping, as previously described (1).

Whole genome sequencing

DNA extraction for long-read sequencing was performed on a Maxwell RSC Instrument (Promega, Madison, USA) using the Maxwell RSC Blood DNA Kit (Promega, Madison, Wisconsin, USA) following the manufacturer’s instructions. Illumina sequencing was performed on a NextSeq-500 system as previously described (2). Nanopore libraries were prepared using Rapid Barcoding Kit V14 (SQK-RBK114.24, Oxford Nanopore Technologies, Oxford, UK) and sequenced on R10.4.0 flow cells (FLO-MIN114) using a Mk1C machine for 72h.

Genome assembly

Illumina *de novo* genome assemblies were obtained as previously described (3) using SPAdes v4.0.0 on preprocessed reads. Nanopore raw data were basecalled with Dorado (v 0.5.3) and assembled with Flye (v 2.9).

Genotyping, MLVA and pertactin analysis

Genotyping was performed using the BIGSdb-Pasteur Bordetella database (<https://bigsdb.pasteur.fr/bordetella/>) using the Bp_vaccine antigen and macrolide resistance typing schemes, as described in Rodrigues *et al* (4). Multi Locus VNTR Analysis (MLVA) was determined using blastN considering data from Schouls *et al* (5). Pertactin production deficiency was investigated using blastN using B1917_2793 as reference and considering the list of all the events that are disrupting the prn expression (6) and Bigsdb alleles nomenclature for locus prn within Autotransporters typing scheme(2).

Whole-genome single nucleotide polymorphisms (SNP) and 23S rRNA gene analyses

SNP calling was performed from Illumina reads mapped against the complete Tohama I reference genome (Accession number: NC_002929), as previously described (6), using genomic data from 2,412 isolates: 1586 from France and 826 from China (723 genomes from the project PRJNA1071282, (7) and 103 from the project PRJNA908268 (8).

Mutations in the 23S-RNA were detected using an in-house script using the bcftools v1.6 mpileup. This allowed to count the reads supporting the mutation A2047G and distinguish isolates with 0, 1, 2 or 3 mutated copies of the 23S-RNA (*i.e.*, coexistence of alleles 1 and 13, or not).

Phylogenetic analysis

The SNP-based alignment was used to infer the phylogenetic relationships among isolates, using IQ-tree v2.3.6 and a GTR+F substitution model (9). The figures were generated using ggtree v3.2 (10). A tree visualisation was performed using Microreact (11), with an interactive project created for open exploration of the dataset (see Figure 1 legend).

qPCR detection of macrolide-resistant *B. pertussis* isolates

A qPCR method was set-up to detect from clinical samples or from extracted DNA, the isolates with the 23S rRNA A2047G mutation conferring macrolide resistance. The method was developed for the LC480 (version II, Roche) thermocycler, using the same primers as a previously described Cycleave Real-Time PCR Assay (12). Briefly, the qPCR targeting *23S rRNA* differentiates two alleles of the 23S subunit ribosomal RNA gene of *Bordetella pertussis* (the gene is present in 3 copies per genome). The A2047G mutation distinguishes macrolide-susceptible isolates, with an A at position 2047, from macrolide-resistant *Bordetella pertussis* isolates (MRBP), with a G at position 2047. Two distinct fluorescent probes were designed targeting the A2047G mutation: one specific for the susceptible allele (**23s_rRNA_SUS**: 6FAM-CGGCTAGACGGAAAGACCCCA-BHQ2) and the other for the resistant allele (**23s_rRNA_RES:** HEX-CGGCTAGACGGGAAGACCCCA-BHQ2). These probes target a 123 bp fragment of the 23S ribosomal RNA of *B. pertussis*; they identify the A2047G mutation after PCR amplification using the following primers: **23S-rRNA-F**, 5'-GAATGGCGTAACGATG-3'; **23S-rRNA-R,** 5'- TGCAAAGCTACAGTAAAGG -3'). Details of the method can be found on protocol.io (13).

The analytical sensitivity of our optimized assay was evaluated using a series of ten-fold dilutions of DNA extracted from *Bordetella pertussis* CIP 112499 (FR4991, the first macrolide-resistant strain identified in France). Each dilution was independently tested in triplicate. Under our conditions, the limit of detection (LOD) was determined to be 10^5 CFU per PCR reaction, corresponding to a Ct value below 32 in the qPCR targeting the pertussis toxin subunit A gene (qPCR PTa). Accordingly, we recommend performing the in-house 23S rRNA qPCR only on DNA extracted from respiratory samples that have previously tested positive for pertussis toxin with a Ct below 30 in the PTa assay, reflecting an adequate bacterial load for reliable genotyping.

**Supplementary Tables**

**Table S1**. Description of the five main genotypes circulating in France in 2024 and their related phenotypes

| **Genotypes** | | **Number of isolates**  **(%)** | **Number of MRBP isolates** | **PT phenotype** | **FHA phenotype** | **PRN phenotype** | **FIM serotype(s)** |
| --- | --- | --- | --- | --- | --- | --- | --- |
| **Bp-agST 4** | ***ptxP3****, ptxA1, ptxB1, ptxC4, ptxD1, ptxE4, fhaB1, fim2-1,* ***fim3-1*** | 242 (69.7%) | **14** | PT+ | FHA+ | 1 PRN+  **13 PRN-neg** | 230 FIM2  11 FIM3  1 nd |
| **Bp-agST 9** | ***ptxP3****, ptxA1, ptxB1, ptxC4, ptxD1, ptxE4, fhaB1, fim2-1,* ***fim3-2*** | 76  (21.9%) | 0 | PT+ | **3 FHA-neg** | 75 PRN+  **1 PRN-neg** | 9 FIM2  60 FIM3  1 FIM2+3  6 nd |
| **Bp-agST 135** | ***ptxP3****, ptxA1, ptxB1, ptxC4, ptxD1, ptxE4,* ***fhaB101****, fim2-1,* ***fim3-1*** | 11  (3.2%) | 0 | PT+ | FHA+ | PRN+ | 11 FIM2 |
| **Bp-agST 93** | ***ptxP1****, ptxA1, ptxB1, ptxC1, ptxD1, ptxE4, fhaB1, fim2-1,* ***fim3-26*** | 11  (3.2%) | 0 | PT+ | FHA+ | PRN+ | 3 FIM2  4 FIM3  3 FIM-neg  1 nd |
| **Bp-agST 34** | ***ptxP1****, ptxA1, ptxB1, ptxC1, ptxD1, ptxE4, fhaB1, fim2-1,* ***fim3-4*** | 7  (2.0%) | 0 | PT+ | FHA+ | PRN+ | 7 FIM2 |

MRBP, macrolide-resistant *B. pertussis*; PT, pertussis toxin; FHA, filamentous hemagglutinin; PRN, pertactin; FIM, fimbriae; nd, not determined**.**

**Table S2**. Characteristics of the 14 macrolide-resistant *B. pertussis* from France, 2024

| **MRBP isolate ID** | ***23S rRNA*** | ***ptxP*** | ***ptxA*** | ***ptxB*** | ***ptxC*** | ***ptxD*** | ***ptxE*** | ***fhaB*-2400_5550** | ***fim2*** | ***fim3*** | **Bp-agST** | **VNTR1** | **VNTR3a** | **VNTR3b** | **VNTR4** | **VNTR5** | **VNTR6** | **MT** | **prn** | **MRBP introductions** |
| --- | --- | --- | --- | --- | --- | --- | --- | --- | --- | --- | --- | --- | --- | --- | --- | --- | --- | --- | --- | --- |
| FR7302 | **13** | 3 | 1 | 1 | 4 | 1 | 4 | 1 | 1 | 1 | **4** | 8 | 7 | 0 | 7 | 6 | 8 | **MT28** | SNP C531T with IS481 inserted | I1 |
| FR7575 | **13** | 3 | 1 | 1 | 4 | 1 | 4 | 1 | 1 | 1 | **4** | 8 | 7 | 0 | 7 | 6 | 8 | **MT28** | SNP C531T with IS481 inserted | I2 |
| FR7673 | **13** | 3 | 1 | 1 | 4 | 1 | 4 | 1 | 1 | 1 | **4** | 8 | 7 | 0 | 7 | 6 | 8 | **MT28** | SNP C531T | I3 |
| FR8119 | **13** | 3 | 1 | 1 | 4 | 1 | 4 | 1 | 1 | 1 | **4** | 8 | 7 | 0 | 7 | 6 | 8 | **MT28** | SNP C531T with IS481 inserted | I2 |
| FR8120 | **13** | 3 | 1 | 1 | 4 | 1 | 4 | 1 | 1 | 1 | **4** | 8 | 7 | 0 | 7 | 6 | 8 | **MT28** | SNP C531T with IS481 inserted | I2 |
| FR8321 | **13** | 3 | 1 | 1 | 4 | 1 | 4 | 1 | 1 | 1 | **4** | 8 | 7 | 0 | 7 | 6 | 8 | **MT28** | SNP C531T with IS481 inserted | I2 |
| FR8322 | **13** | 3 | 1 | 1 | 4 | 1 | 4 | 1 | 1 | 1 | **4** | 8 | 7 | 0 | 7 | 6 | 8 | **MT28** | SNP C531T with IS481 inserted | I2 |
| FR8344 | **13** | 3 | 1 | 1 | 4 | 1 | 4 | 1 | 1 | 1 | **4** | 8 | 7 | 0 | 7 | 6 | 10 | **MT30** | SNP C531T with IS481 inserted | I2 |
| FR8384 | **13** | 3 | 1 | 1 | 4 | 1 | 4 | 1 | 1 | 1 | **4** | 8 | 7 | 0 | 7 | 6 | 8 | **MT28** | SNP C531T with IS481 inserted | I1 |
| FR8447 | **13** | 3 | 1 | 1 | 4 | 1 | 4 | 1 | 1 | 1 | **4** | 8 | 7 | 0 | 7 | 6 | 8 | **MT28** | SNP C531T with IS481 inserted | I2 |
| FR8572 | **13** | 3 | 1 | 1 | 4 | 1 | 4 | 1 | 1 | 1 | **4** | 8 | 7 | 0 | 7 | 6 | 8 | **MT28** | SNP C531T with IS481 inserted | I2 |
| FR8573 | **13** | 3 | 1 | 1 | 4 | 1 | 4 | 1 | 1 | 1 | **4** | 8 | 7 | 0 | 7 | 6 | 8 | **MT28** | SNP C531T with IS481 inserted | I2 |
| FR8574 | **13** | 3 | 1 | 1 | 4 | 1 | 4 | 1 | 1 | 1 | **4** | 8 | 7 | 0 | 7 | 6 | 8 | **MT28** | SNP C531T with IS481 inserted | I2 |
| FR8581 | **13** | 3 | 1 | 1 | 4 | 1 | 4 | 1 | 1 | 1 | **4** | 8 | 7 | 0 | 7 | 6 | 8 | **MT28** | SNP C531T with IS481 inserted | I2 |

MT: MLVA type; prn: pertactin gene; The integers correspond to the allele numbers of the indicated genes or VNTR loci.
